# SeqFirst: Building equity access to a precise genetic diagnosis in critically ill newborns

**DOI:** 10.1101/2024.09.30.24314516

**Authors:** Tara L. Wenger, Abbey Scott, Lukas Kruidenier, Megan Sikes, Alexandra Keefe, Kati J. Buckingham, Colby T. Marvin, Kathryn M. Shively, Tamara Bacus, Olivia M. Sommerland, Kailyn Anderson, Heidi Gildersleeve, Chayna J. Davis, Jamie Love-Nichols, Katherine E. MacDuffie, Danny E. Miller, Joon-Ho Yu, Amy Snook, Britt Johnson, David L. Veenstra, Julia Parish-Morris, Kirsty McWalter, Kyle Retterer, Deborah Copenheaver, Bethany Friedman, Jane Juusola, Erin Ryan, Renee Varga, Dan Doherty, Katrina Dipple, Jessica X. Chong, Paul Kruszka, Michael J. Bamshad

## Abstract

Access to a precise genetic diagnosis (PrGD) in critically ill newborns is limited and inequitable because complex inclusion criteria used to prioritize testing eligibility omits many patients at high risk for a genetic condition. SeqFirst-neo is a program to test whether a genotype-driven workflow using simple, broad exclusion criteria to assess eligibility for rapid whole genome sequencing (rWGS) increases access to a PrGD in critically ill newborns. All 408 newborns admitted to a neonatal intensive care unit between January 2021 and February 2022 were assessed and of 240 eligible infants, 126 were offered rWGS (i.e., intervention group [IG]) and compared to 114 infants who received conventional care in parallel (i.e., conventional care group [CCG]). A PrGD was made in 62/126 (49.2%) IG neonates compared to 11/114 (9.7%) CCG infants. The odds of receiving a PrGD was ∼9 times greater in the IG vs. the CCG, and this difference was maintained at 12 months follow up. Access to a PrGD in the IG versus CCG differed significantly between infants identified as non-white (34/74, 45.9% vs. 6/29, 20.7%; p=0.024) and Black (8/10, 80.0% vs. 0/4; p=0.015). Neonatologists were significantly less successful at predicting a PrGD in non-white than non-Hispanic white patients. Use of a standard workflow in the IG with a PrGD revealed that a PrGD would have been missed in 26/62 (42%) of infants. Use of simple, broad exclusion criteria that increases access to genetic testing significantly increases access to a PrGD, improves access equity and results in fewer missed diagnoses.

## Introduction

Knowledge of the genetic basis of pediatric diseases has increased exponentially over the past decade and genetic variation plays a role in virtually all pediatric conditions. New technologies to rapidly and inexpensively interrogate human genomes, powerful computational approaches to identify disease-causing (i.e., pathogenic) variants, and high-throughput functional assays to facilitate development of precision therapeutics, have converged to create an unprecedented opportunity to capitalize on this information and use genomics to revolutionize pediatric healthcare. Indeed, the widespread application of genomics (e.g., exome sequencing / whole genome sequencing [ES / WGS]) in clinical genetics has had a major impact on the ability to make a precise genetic diagnosis (PrGD) in patients with rare conditions (RCs).^1,2^ Use of ES / WGS increases diagnostic rates, enables faster diagnosis, reduces costs, and improves both family and provider satisfaction while a delayed or absent PrGD can result in both missed or inappropriate interventions.^3–21^ Nevertheless, integration of ES / WGS into most pediatric clinical programs and specialty services has been modest at best.^22–29^

Rapid ES / WGS is transforming diagnosis and care of RCs in critically ill newborns. Evidence from more than forty studies that collectively have evaluated >3,500 families is compelling.^3–13,15–21,26,29–54^ Yet, the impact of the widespread use of rapid ES / WGS has been blunted, to date, by the lack of availability of effective service-delivery models that support scalability (i.e., widespread adoption in neonatal intensive care units [NICUs], offering testing to all infants with a high prior risk for a genetic condition, etc.) limiting equitable access to a PrGD (i.e., offering a genetic test and receipt of a test result that explains a patient’s clinical findings).^55–57^ Indeed, most children in the United States who could benefit from a PrGD aren’t offered advanced genetic testing since the availability of genetic testing is highly dependent upon institution, geography, and social class.^6^ Another challenge is that non-specific presentations of RCs are common, and thus suspicion of a RC as a prerequisite to request genetic testing excludes many, if not most, individuals with a RC from access to testing.

Moreover, the ability to offer testing is most compromised in communities that have traditionally been underserved and disproportionately represented by families that identify as Black, Indigenous, People Of Color and / or living in rural areas.^58,59^ This disparity is compounded by structural racism resulting in inequities in NICU care for underrepresented minorities. These inequities are driven in part by a lower likelihood of admission to a high quality medical center and differences in care compared to white infants admitted to the same NICUs.^60–66^ This is a major gap in healthcare.

SeqFirst is a research initiative established to develop and test innovative genotype-driven service delivery models in pediatric care settings that serve diverse communities with varied levels of infrastructure for providing clinical genetic services. It currently includes three arms: SeqFirst-Ddi, focused on children under three years of age newly found by their parents or primary care providers to have atypical development; SeqFirst-All Kids Included, aimed at supporting access to a PrGD in underserved communities; and SeqFirst-neo, a project to develop and test approaches to center equity for a PrGD at the initial point of care of infants with a critical illness. Each of these arms also represent an opportunity to develop and deploy complementary technological strategies to support provider readiness (e.g., telemedicine consults, virtual consenting, self-guided return of results, etc.).

Herein we present the results of Phase 1 of SeqFirst-neo in which we implemented a genotype-driven workflow using simple, broad exclusion criteria to assess eligibility for rapid whole genome sequencing (rWGS) and tested its impact on several outcomes including access to a PrGD, diagnostic yield, and changes in clinical management in critically ill newborns. We also compared these outcomes among groups delimited by self or provider assigned population descriptors including race to test whether access to a PrGD was equitable.

### Subject, material, and methods

#### Patient selection and enrollment

This study was reviewed and approved by the University of Washington IRB # STUDY00008810 and consent was obtained for each participant. Exclusion criteria were developed by a multidisciplinary team of care-providers and researchers including clinical genetics, genetic counseling, and neonatology. Exclusion criteria included a corrected age > 6 months or clinical findings fully explained by physical trauma, infection, or complications of prematurity. Infants with clinical findings initially considered fully explained by physical trauma, infection, or complications of prematurity who developed, as judged by a neonatologist, an atypical clinical course (e.g., excessive bleeding) during their hospitalization were offered enrollment. Infants with a pre-existing PrGD via prenatal genetic testing or postnatal testing at their birth hospital were also excluded. Eligibility did not require sample collection from any biological parents.

From January 2021 to February 2022, the admission notes of each infant admitted to the NICU at Seattle Children’s Hospital (SCH), a tertiary pediatric hospital located in an urban setting, were briefly (i.e., 2-3 minutes) assessed remotely each day by a clinical geneticist (T.W., K.D., M.B.) to determine eligibility (Figure 1). This assessment was then confirmed with the neonatologist on service. Eligible families were approached by a research team member, either a genetic counselor or clinical geneticist, to assess parental interest in enrollment. Interpreter services were used as needed. Infants enrolled were assigned to the intervention group (IG). Conventional clinical care and testing, including genetic testing, continued in parallel for each infant enrolled in SeqFirst without restriction on tests ordered. Families were given the option to receive secondary findings. If no clinical genetics consult had been obtained through conventional clinical care, reporting of a variant of unknown significance, likely pathogenic, pathogenic, or secondary result triggered consultation.

**Figure 1.**
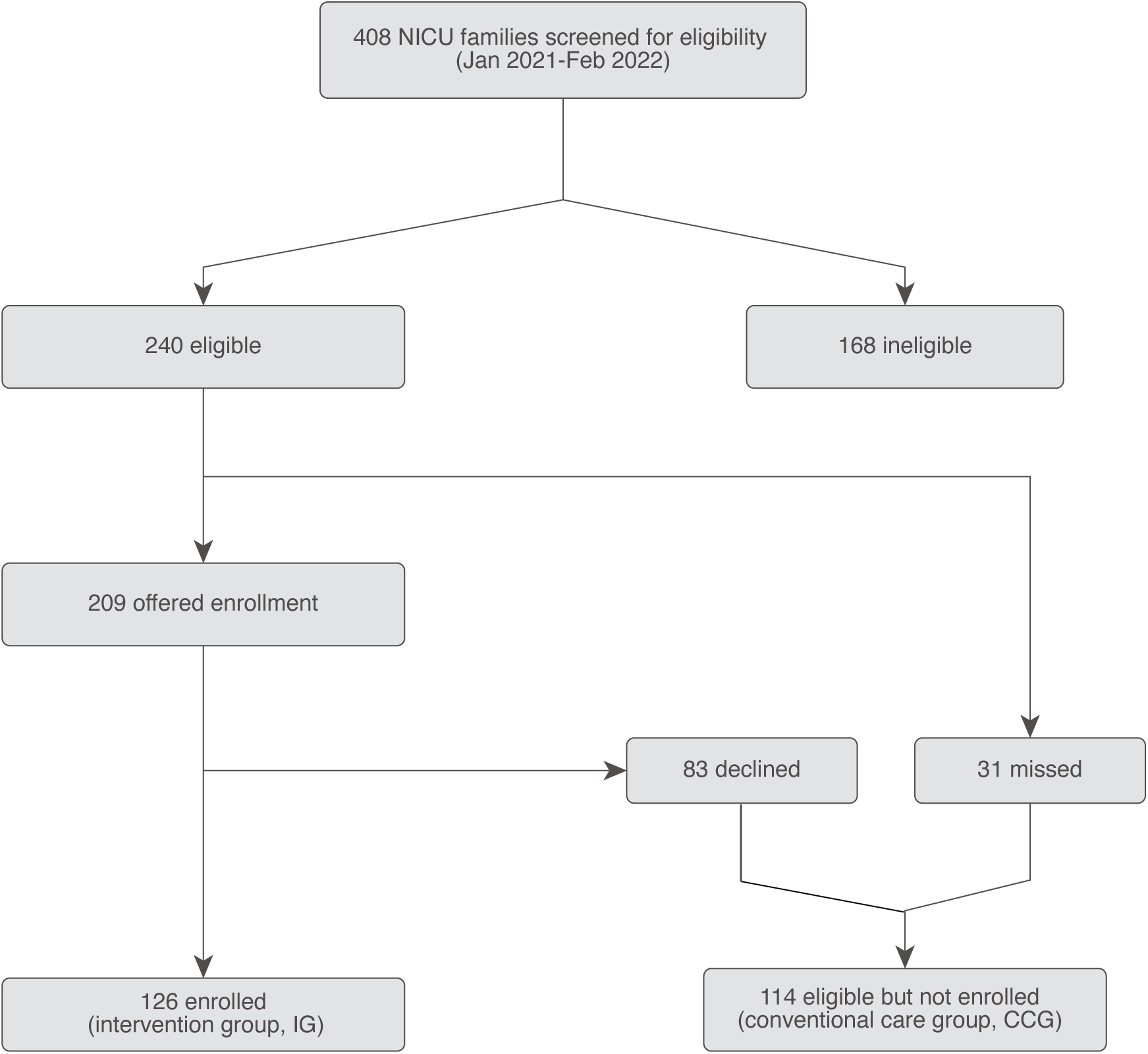
STROBE diagram. SeqFirst-neo cohort ascertainment of interventional group (IG) and conventional care group (CCG) depicted using a STROBE diagram.

Patients who were eligible but not enrolled (e.g., families not available for consent or declined to participate) were assigned to a conventional care group (CCG). Standard clinical care and testing, including genetic testing, continued for each infant in the CCG. The SeqFirst and CCG were well-matched for gestational age at birth, age at admission, sex assigned at birth, admitting diagnosis, and parent- or provider-assigned racial construct (PPARC) (Table 1).

**Table 1:**
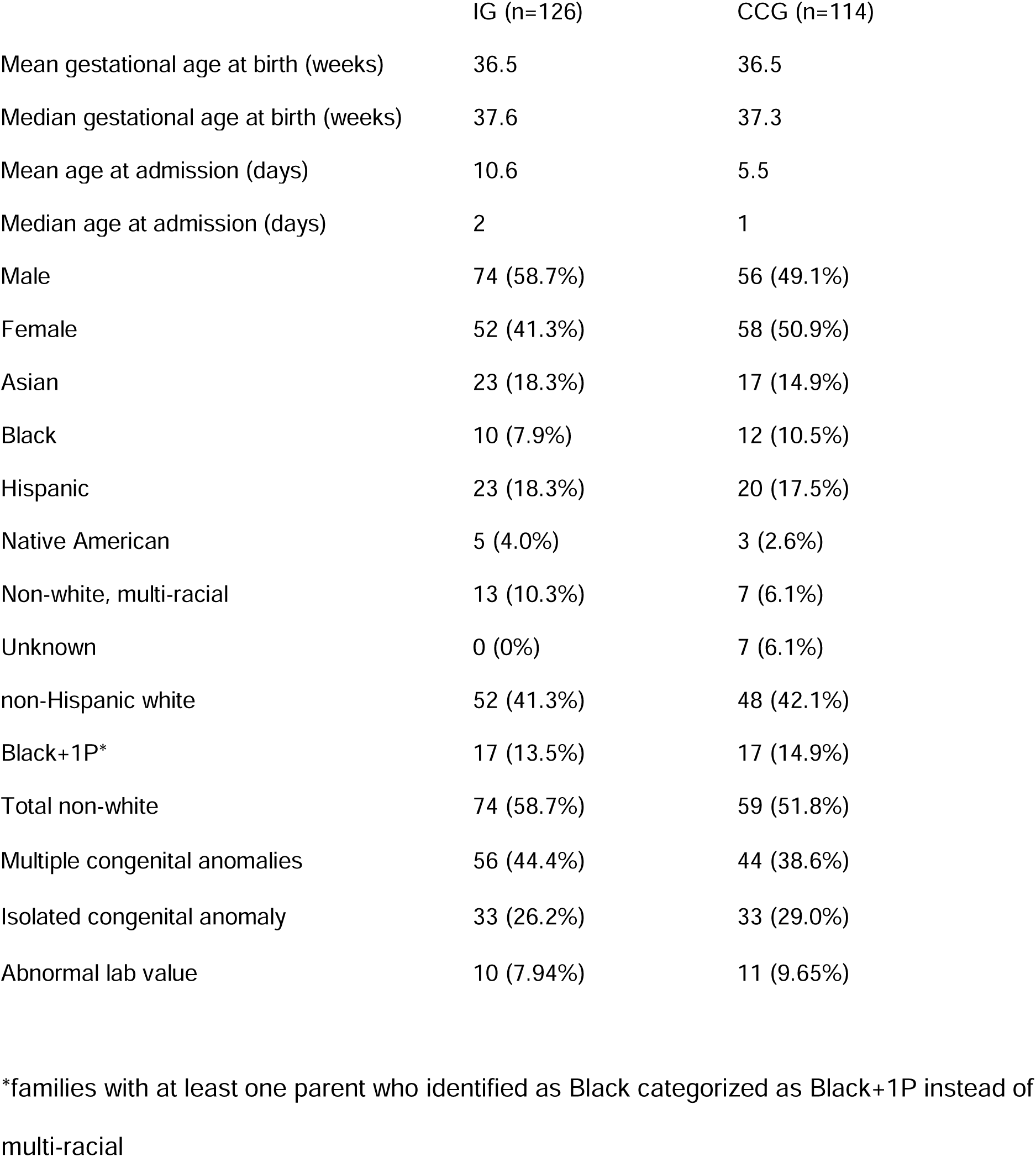
Characteristics of Intervention Group (IG) and Conventional Care Groups (CCG)

#### Rapid whole genome sequencing

Whole blood was collected from the proband, parents, and, if necessary, additional family members (e.g., siblings). rWGS was performed using a CLIA-certified, commercially available clinical test (*GenomeXpress*) offered by GeneDx. In brief, PCR-free whole genome sequencing libraries were prepared using Illumina DNA PCR-Free Prep, Tagmentation kit following the manufacturer’s protocol (Illumina Inc.). Massively parallel (NextGen) sequencing was performed on an Illumina platform. Average mean sequencing coverage was at least 40x across the genome, with a minimum threshold of 30x for any proband sample. Bi-directional sequence reads were assembled and aligned using Dragen to reference sequences based on NCBI RefSeq transcripts and human genome build GRCh37/UCSC hg19. Using a custom-developed analysis tool, data were filtered and analyzed to identify sequence variants, repeat expansions in *FMR1* and *DMPK*, homozygous loss of *SMN1* exon 8, and most deletions and duplications greater than 1 kb in size. PCR-free whole genome sequencing libraries were prepared using Illumina DNA PCR-Free Prep, Tagmentation kit following the manufacturer’s protocol (Illumina Inc.). Massively parallel (NextGen) sequencing was performed on an Illumina platform. Average mean sequencing coverage was at least 40x across the genome, with a minimum threshold of 30x for any proband sample. Bi-directional sequence reads were assembled and aligned using Dragen to reference sequences based on NCBI RefSeq transcripts and human genome build GRCh37/UCSC hg19. Using a custom-developed analysis tool, data were filtered and analyzed to identify sequence variants, repeat expansions in *FMR1* and *DMPK*, homozygous loss of *SMN1* exon 8, and most deletions and duplications greater than 1 kb in size. Additional sequencing technology and variant interpretation protocol has been previously described^67^. The general assertion criteria for variant classification are publicly available on the GeneDx ClinVar submission page (https://www.ncbi.nlm.nih.gov/clinvar/docs/review_status/#ac). All reported variants were confirmed by an orthogonal method in the proband and parents, if available. A result including nuclear pathogenic and/or likely pathogenic variants potentially explaining a patient’s clinical findings was verbally communicated to the research team within seven calendar days and written results within fourteen days.

Each reported variant was interpreted by the research team including at least two clinical geneticists and a genetic counselor and each reported variant(s)/gene(s) was assigned to one of five categories: clinical findings explained, clinical findings likely explained, clinical findings partially explained (i.e., a variant(s) explains at least one but not all clinical findings), clinical findings not explained, secondary finding unrelated to clinical findings. Assignments were made based on a combination of clinical judgement of phenotype specificity and similarity to previously reported gene-disease phenotypes, use of the ClinGen gene-disease validity framework, and use of the ACMG/AMP sequence variant classification (SVC) and ACMG/ClinGen CNV classification guidelines (plus soon to be released ACMG/AMP/CAP/ClinGen v4 SVC guidelines). We required at least a moderate level of gene-disease validity for a gene to be potentially considered explanatory and at least near-moderate for likely explained. A PrGD was assigned to an infant if they had at least one variant categorized as clinical findings explained, likely explained, or partially explained.

#### Return of results and collection of clinical data

Upon verbal receipt of a result, the research team communicated the result to the clinical team. Results that were considered explanatory (i.e., explained, likely explained, or partially explained) and secondary results were returned to the family by the clinical genetics consultants. Results that were non-explanatory were returned to the family by the research team.

Information about the clinical course of each infant in the IG was collected from the electronic medical record (EMR). Additional information was collected via electronic surveys administered to parents/guardians upon completion of consent (pregnancy, birth, and family history), after results disclosure (impact of results returned), and at 6, 12, and 18 months (developmental milestones and healthcare utilization) after enrollment. For CCG infants, clinical data including gestational age at birth, age at admission, diagnoses at admission, and PrGD were collected from the EMR.

Access to a PrGD was assessed upon receipt of results, and at 3 and 15 months after enrollment. These latter two time points were selected to allow for completion of additional testing (e.g., orthogonal validation of results, evaluation for somatic mosaicism, etc.) and clinical evaluations, both of which were used to assist adjudication of variants found via rWGS and identify variants not detected by rWGS. Changes in management were defined as actions taken as a result of a PrGD including additional laboratory testing, imaging, consultations, procedures, and changes in medical treatment.

Race and ethnicity information were collected by review of family history obtained by a genetic counselor or clinical geneticist, and from race and ethnicity terms reported in the EMR. Information collected by a provider was prioritized over that obtained from the EMR. Each individual was assigned by the research team to one of six PPARC: Asian, Black, Hispanic, Native American, non-Hispanic white and non-white multi-racial (NWMR).

#### Outcomes and Statistical analysis

Primary outcomes assessed were: 1) access to a PrGD defined as the percentage of patients eligible for testing who received a PrGD; 2) diagnostic yield defined in the IG as the percentage of patients tested with a variant identified by rWGS that likely explained, explained, or partially explained the clinical findings of a patient and in the CCG as a variant identified by any genetic testing that likely explained, explained, or partially explained the clinical findings of a patient; 3) access to a PrGD and diagnostic yield among PPARC groups; and 4) PrGD missed in the IG defined as families with a PrGD in whom genetic testing was not offered as part of conventional care. In the IG, access to a PrGD and diagnostic yield are identical metrics as all patients were tested.

The primary comparison between rate of PrGD diagnosis in the IG vs. CCG was made using a generalized linear model (logistic regression), with χ^2^ or Fisher’s exact test applied to smaller subsample comparisons. Fisher’s exact test was used when greater than 20% of cells had fewer than five individuals. Regressions were conducted in R version 4.4.1^68^, with condition coded as CCG=0 and IG=1. A p-value of 0.05 or less was deemed statistically significant.

## Results

From January 2021 to February 2022, 408 infants were admitted to the NICU (Figure 1). A total of 168 patients were ineligible (Supplementary Table 1) because their clinical findings were explained by birth trauma (n=64), prematurity (n=47), infection (n=36), a pre-existing PrGD (n=15) and “other” reasons including observation for a condition not identified (n=6). Two hundred forty infants were eligible to participate. Thirty-one infants were discharged, transferred from the NICU, or died before their families were available for eligibility screening and were therefore considered “missed.” Two hundred nine families were screened, and 126 (60%) enrolled in the IG. Most enrolled patients (n=89, 71%) had either multiple congenital anomalies (n=56, 44%) or an anomaly of a single organ / body part (n=33, 26%). Twelve infants had either abnormal laboratory tests (n=10, 8%) or hematological abnormalities (n=2, 2%); 9 (7%) had seizures, 6 (5%) had single or multi-organ failure, 3 (2%) had strokes, and 7 (6%) had other unclassified admission diagnoses (e.g., congenital ichthyosis, meconium ileus, vocal cord paresis, etc. [Supplementary Table 2]).

A total of 83 families declined to participate (Supplementary Table 3). Thirty-two parents did not respond to requests for interview. The remaining families cited being overwhelmed (n=26), concerns that testing was unnecessary (n=13), discomfort with research (n=7), concerns over privacy (n=3), or concerns about phlebotomy (n=2). Infants who were eligible but “missed” (n=31) or whose parents declined to participate (n=83) were assigned to the CCG (n=114).

Eighty-five infants had one or more variants or events reported by GeneDx that disrupted 120 genes, genomic regions or the mitochondrial genome (Figure 2). No variants were reported in 41 infants (Figure 2). Clinical interpretation of these variants and events led to a rapid PrGD in a total of 62/126 (49%) infants in the IG in the first 90 days post enrollment (Table 2, Figure 3). Most infants were reported to have variant(s) that explained (n=37; 29%) or likely explained (n=21; 17%) their clinical findings (Figure 2). Clinical findings were partially explained in 4 (3%) infants. No PrGD was made in 64/126 infants (51%) including 7 (6%) who received only secondary findings and 57 (45%) in whom no explanatory variants were identified. In the CCG, a PrGD was made in 11/114 (10%) infants within their first 90 days of hospitalization, significantly fewer (49% vs. 10%, p<0.00001) than in the IG (Figure 3). This difference was sustained over the ensuing 12 months, 63/126 or 50%, in the IG compared to 12/114 or 11% in the CCG (p<0.00001). This difference reflected differences in access to genetic testing as only a subset (n=46) of infants in the CCG underwent clinical genetic testing, and even fewer (n=7) underwent ES in the first fifteen months of life (Supplementary Table 4). But even among patients who received any type of genetic testing, the difference in diagnostic yield in PrGD between the IG and the CCG was significant at 90 days (62/126, 49% vs. 11/45, 24%; 0.0039) and remained significant (63/126, 50% vs. 12/46, 26%; p=0.0087).

**Figure 2.**
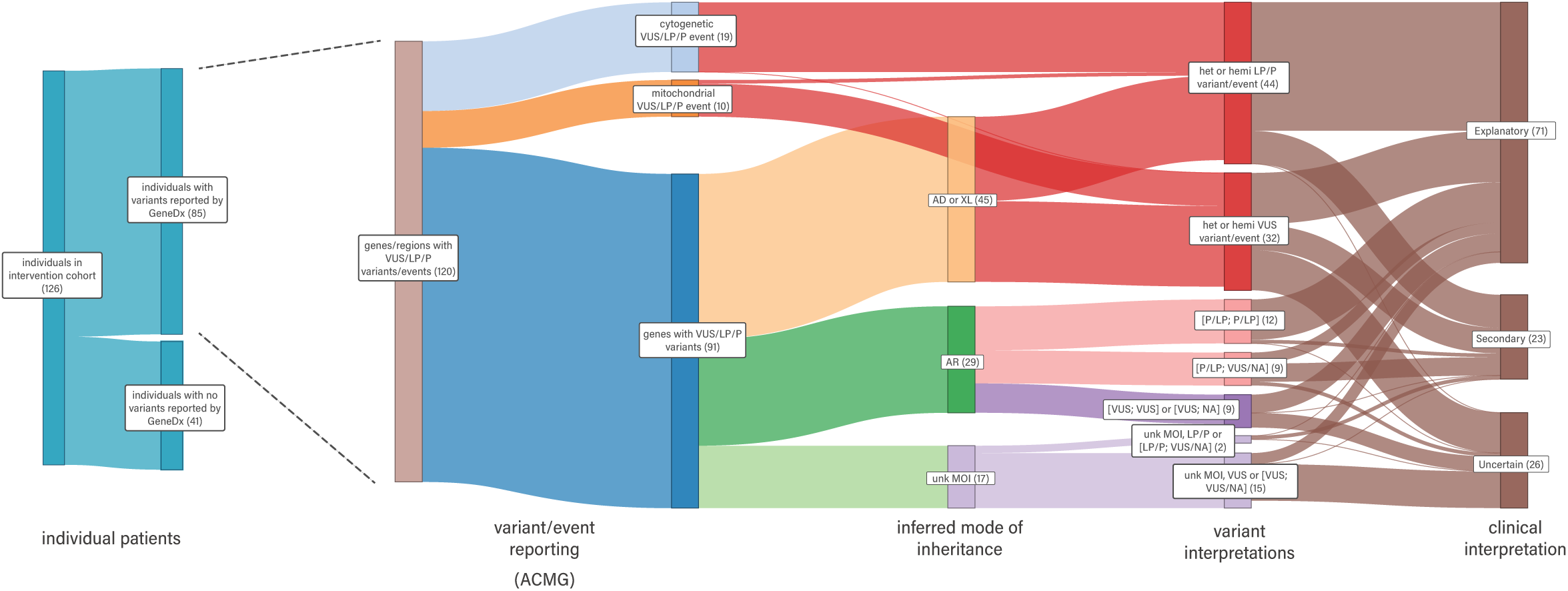
Adjudication of variants reported in patients in the intervention cohort. Of the 126 participants, 85 had one or more potentially explanatory variants reported by GeneDx based on the American College of Medical Geneics and Genomics (ACMG) guidelines for clinical sequence interpretation (variant of uncertain significance [VUS], likely pathogenic [LP], pathogenic [P]). We grouped these variants by the genes/regions in which they were reported (i.e., single gene, cytogenetic event, mitochondrial event) and by the inferred inheritance pattern(s) of the condition(s) they underlie (autosomal dominant [AD] or X-linked [XL], autosomal recessive [AR], or unknown mode of inheritance [MOI] for genes that underlie both AD and AR conditions. Variant(s) or event(s) and the MOI were then adjudicated together to determine whether each combination was explanatory of clinical findings (either partly or fully), unrelated to clinical findings and secondary, or of uncertain relationship to clinical findings of each participant. Variants / events in each participant were counted separately so for example the same genotype in two participants would be counted twice.

**Figure 3.**
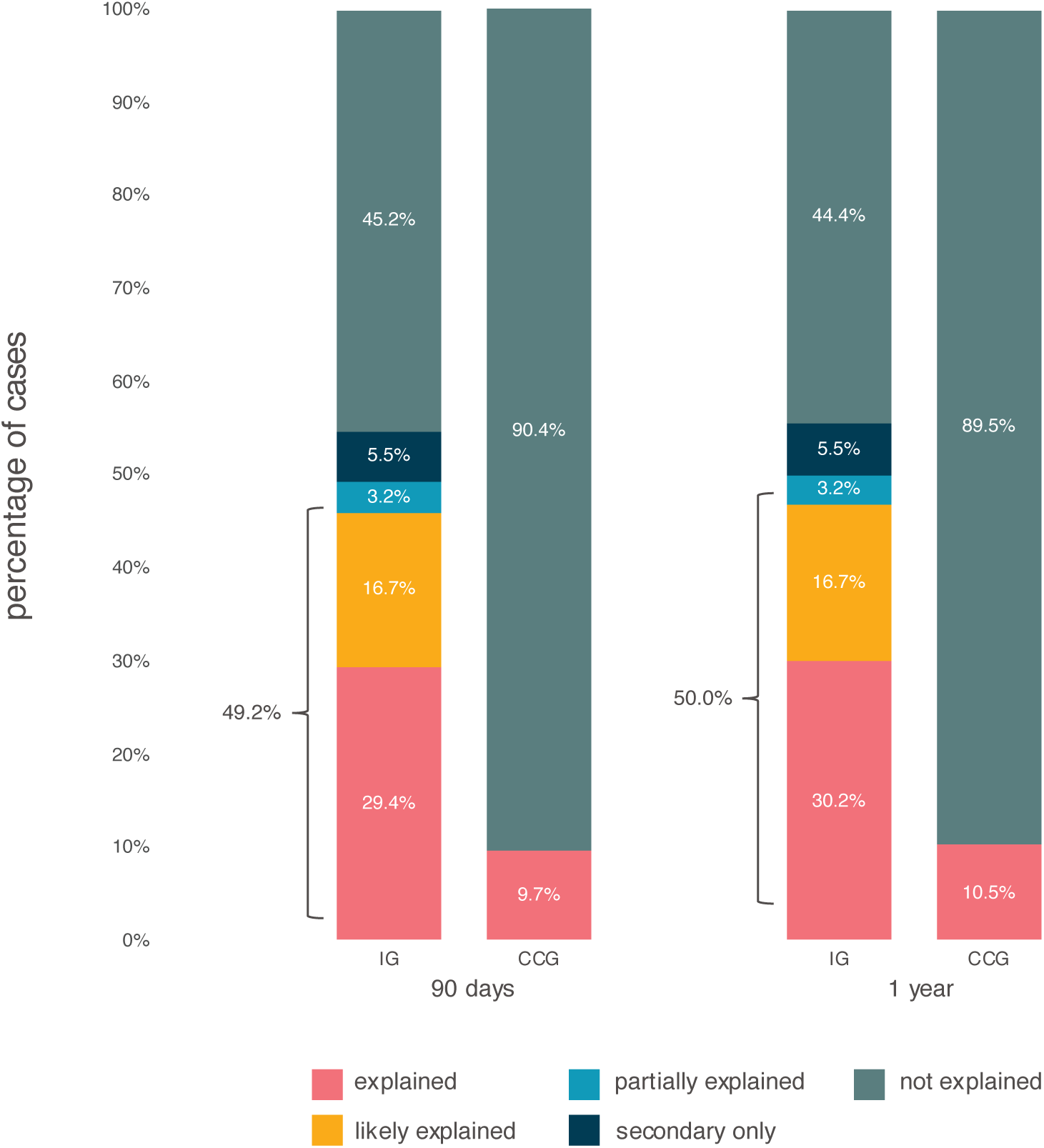
Diagnostic yield in the intervention group. Overall diagnostic yield stratified by test result category in the intervention group (IG) and the conventional care group (CCG) at 90 days and one year post ascertainment.

**Table 2:** Access to a PrGD among different PPARC groups at 90 days.

| PPARC groups | PrGD at 90 days |  |  | p-values |  |
| --- | --- | --- | --- | --- | --- |
|  | Intervention<br>(n=126) | CCG<br>(n=107*) | CCG any<br>testing (n=46) | Intervention vs.<br>CCG | Intervention vs.<br>CCG any<br>testing |
| Asian | 12/23 (52.2%) | 4/17 (23.5%) | 4/8 (50%) | 0.1043 | 0.4759 |
| Black | 8/10 (80%) | 0/12 (0.0%) | 0/4 (0%) | 0.00001 | 0.015 |
| Hispanic | 8/23 (34.8%) | 2/20 (10.0%) | 2/10 (20%) | 0.0764 | 0.6822 |
| Native American | 1/5 (20%) | 0/3 (0.0%) | 0/0 (0%) | 1.0000 | n/a |
| non-white, multi-racial | 5/13 (38.5%) | 0/7 (0.0%) | 0/7 (0%) | 0.1137 | 0.1137 |
| non-Hispanic white | 28/52 (53.8%) | 5/48 (10.4%) | 5/17 (10.4%) | <0.00001 | 0.0988 |
| total | 62/126 (49.2%) | 11/107 (10.3%) | 11/46 (23.9%) | <0.00001 | 0.0031 |
| total non-white | 34/74 (45.9%) | 6/59 (10.2%) | 6/29 (20.7%) | <0.00001 | 0.0241 |
| Black+1P | 11/17 (45.9%) | 0/17 (0.0%) | 0/8 (0%) | 0.0001 | 0.0029 |

\*CCG cohort denominator for this analysis totals 107 because seven cases had unknown PPARC.

Logistic regression revealed a significant effect of use of exclusion criteria in the IG vs. CCG on the likelihood of receiving a PrGD (β = 2.07, 95% CI = 1.40-2.80, Z=5.84, *p*<.0001). This effect translates to an odds ratio (OR) of 7.92, meaning that infants in the IG were nearly 8 times more likely to receive a PrGD than infants in the CCG group, and the odds of receiving a PrGD increased by 692% for infants in the IG vs. CCG group. This effect remained significant after controlling for infant sex and race/ethnicity in the model (β = 2.21, 95% CI = 1.53-2.98, Z=6.02, *p*<.0001, OR=9.16).

Of 126 patients in the IG, the diagnostic yield varied by phenotypic categorial group (Table 3, Figure 4A) from 33% in those with isolated structural anomalies to 70% in infants with laboratory abnormalities and infants with clinical findings not easily categorized (71%) (Supplementary Table 5). The diagnostic yields in infants with multiple congenital anomalies (MCA; 54%), whether with (57%) or without (50%) congenital heart defects (CHD), and seizures (56%) were similar. A PrGD was made in both patients with hematological abnormalities but none of the 3 patients with stroke. A total of 68 conditions were identified in the 63 infants who received a PrGD including 59 infants with a single condition, 3 infants with two conditions, and one infant with three conditions. Inherited or *de novo* autosomal dominant events were responsible for 32 of 68 (47%) conditions found in 31 infants. Twenty-two autosomal recessive conditions were identified in 21 infants and 6 X-linked conditions were identified in 6 infants. Eleven chromosomal abnormalities including aneuploidy, uniparental disomy, microdeletions, or duplications were observed in 11 infants. Four infants had mitochondrial disorders, two as a result of mtDNA mutations and two with autosomal recessive conditions (Supplementary Table 6).

**Figure 4.**
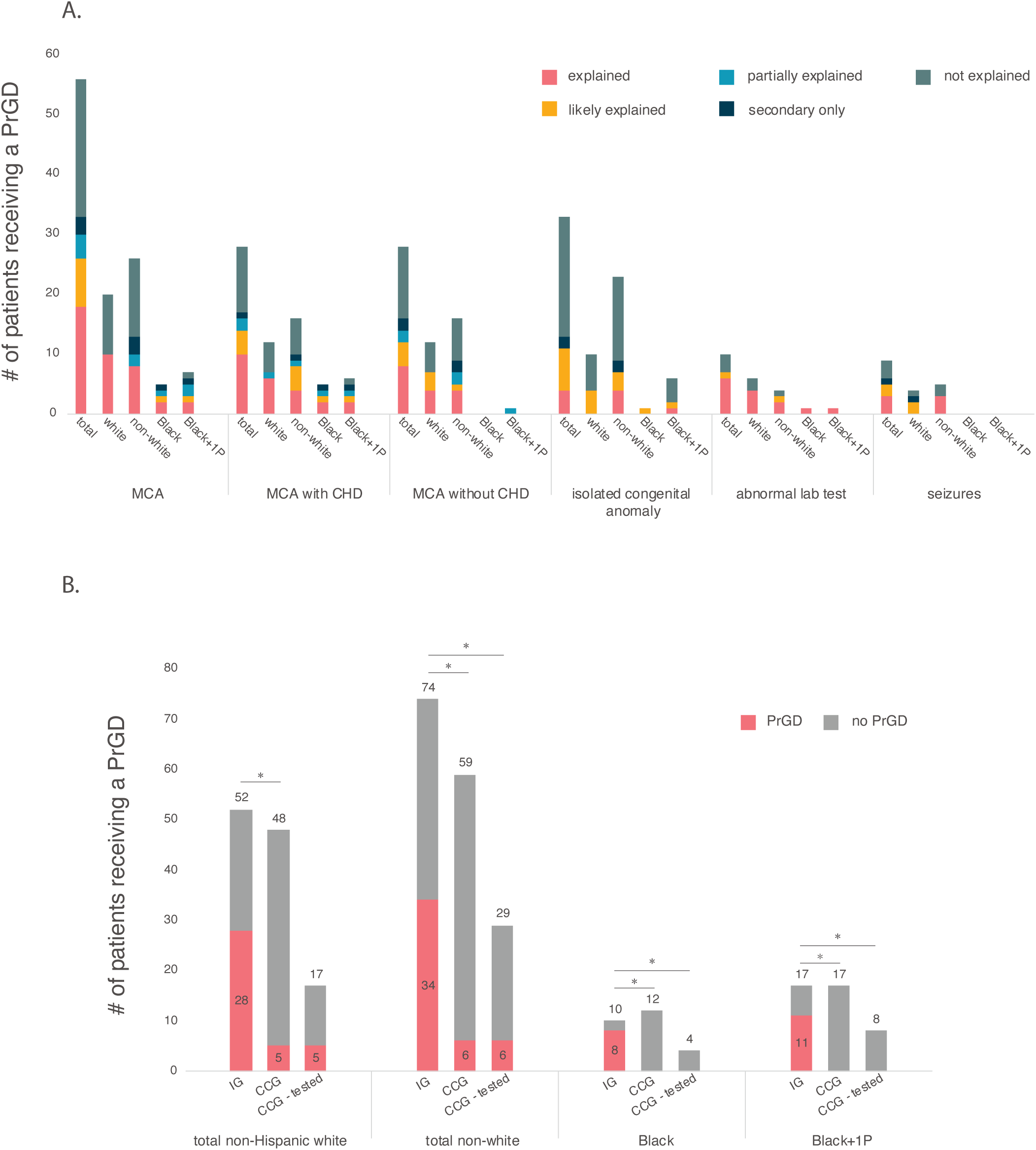
Diagnostic yield in the intervention group by phenotypic presentation and by racial construct. (**A**) Diagnostic yield in the intervention group (IG) stratified by phenotype presentation, parent or provider assigned racial construct (PPARC) and test result category. Abbreviations: multiple congenital anomalies, MCA; congenital heart defects, CHD. (**B**) Access to a PrGD / diagnostic yield in the intervention group (IG), access to a PrGD in the conventional care group (CCG) and the diagnostic yield in the conventional care group who received genetic testing (CCG-tested) stratified by PPARC. Abbreviations: precise genetic diagnosis (PrGD). Asterisks denote statistically significant differences between groups.

**Table 3:** PrGD for Intervention Group by Categorical Admission Diagnosis.

|  | total | explained | Likely<br>explained | partially<br>explained | secondary<br>only | not<br>explained |
| --- | --- | --- | --- | --- | --- | --- |
| MCA | 56 | 18 | 8 | 4 | 3 | 23 |
| MCA w/ CHD | 28 | 10 | 4 | 2 | 1 | 11 |
| MCA w/o CHD | 28 | 8 | 4 | 2 | 2 | 12 |
| Isolated congenital anomaly | 33 | 4 | 7 | 0 | 2 | 20 |
| abnormal lab test | 10 | 6 | 1 | 0 | 0 | 3 |
| hematological abnormality | 2 | 2 | 0 | 0 | 0 | 0 |
| seizures | 9 | 3 | 2 | 0 | 1 | 3 |
| single or multi-organ failure | 6 | 1 | 1 | 0 | 0 | 4 |

|  |  |  |  |  |  |  |
| --- | --- | --- | --- | --- | --- | --- |
| strokes | 3 | 0 | 0 | 0 | 0 | 3 |
| other | 7 | 3 | 2 | 0 | 0 | 2 |

The number of infants with CHDs in the IG is notably lower than is typical of infants admitted to a NICU as most newborns admitted to SCH with CHDs are instead admitted to a Cardiac Intensive Care Unit (CICU). For this reason, from August 2021 to December 2021, we applied the same eligibility criteria and assessment workflow to infants admitted to the CICU until a sample size of 25 families was reached. A total of 52 infants were assessed, 42 of whom were eligible. Forty-one families were interviewed, and 25 (61%) enrolled in SeqFirst. Most enrolled patients (n=15, 60%) had an isolated CHD (Supplementary Table 7). Fourteen of these were simplex families and one infant had a family history of dominantly inherited CHD. Nine infants had CHD as one of multiple congenital anomalies, and one infant was admitted with supraventricular tachycardia. A PrGD was made in a total of 8/25 (32%) infants in the CICU group, 3 /15 (20%) of the infants with isolated CHD, and 5/9 (56%) of the infants with MCA.

### Stratification by patient or provider assigned racial construct (PPARC)

Access to a PrGD among PPARC groups in the IG in the first 90 days of hospitalization ranged between 20% (1/5; Native American) and 80% (8/10; Black) but none of these differences were statistically significant (Table 2, Figure 4B). Access to a PrGD was not statistically different among PPARC groups 12 months later (p=0.1107). Across PPARC groups in the CCG, a PrGD was made in 5 families that identified as non-Hispanic white, 4 that identified as Asian and 2 that identified as Hispanic in the first 90 days. A PrGD was not made in any family that identified as Black, multi-racial, or Native American. A PrGD was made in only one additional infant, (i.e., 12 / 114 families instead of 11 / 114 families) 12 months later. The small number of PrGD in CCG families precluded a robust test of statistical significance of differences across PPARC groups at either assessment point. In both the IG and the CCG, collapsing PPARC assignments and comparing non-Hispanic whites to non-whites or comparing non-Hispanic whites to Black+1P where families with at least one parent who identified as Black were categorized as Black instead of multi-racial did not reveal statistically significant differences in access to a PrGD.

Access to a PrGD in PPARC groups in the IG compared to the CCG at 90 days differed significantly between non-Hispanic white (28/52, 54% vs. 5/48, 10%; p<0.00001), non-white (34/74, 46% vs. 6/59, 10%; p<0.00001), Black (8/10, 80% vs. 0/12; p=0.00001) and Black+1P (11/17, 65% vs. 0/17; p=0.0001) groups. Access to a PrGD did not differ significantly between Asian (p=0.1043), Hispanic (p=0.0764), Native American (p=1.0), or multi-racial (p=0.1137) PPARC groups in the IG compared to the CCG groups. These differences in access to a PrGD between the IG and CCG result in part from differences in access to genetic testing, which differed significantly between the groups. Accordingly, we repeated the analysis but included only CCG infants who were tested. The difference in access to a PrGD in the IG versus CCG non-Hispanic white group was no longer statistically significant (p< 0.1) whereas the difference in access to a PrGD in the IG versus CCG remained statistically significant between non-white (34/74, 46% vs. 6/29, 21%; p=0.024), Black+1P (11/17, 65% vs. 0/8; p=0.003), and Black (8/10, 80% vs. 0/4; p=0.015) groups.

### Yield of precise genetic diagnoses in newborns not suspected of having a genetic condition

For participants in the IG who had a PrGD made in the first 90 days, we compared their outcome to that from testing provided in parallel per a standard clinical workflow. This revealed use of a standard workflow for ascertainment and testing would have been missed a PrGD in 26/62 (42%) of these infants (Supplementary Table 8, Supplementary Figure 1). Nearly one quarter (15/62; 24%) of the infants who received a PrGD in the IG in the first 90 days did not have documented suspicion of a genetic condition in their EMR and therefore were not offered clinical genetics consultation or genetic testing as part of their conventional care. All but one (14/15) of these infants were diagnosed with genetic conditions that do not have recognizable physical features, and most of these infants were described as having an unexpected response to therapies or severity of illness disproportionate to what was expected based upon provisional clinical diagnosis. Moreover, 10/15 (67%) of these infants were identified as a PPARC other than non-Hispanic white. An additional 11/26 infants did not receive a PrGD despite clinical genetics consultation and targeted genetic testing (n=9) or because of lack of access to testing due to demise shortly after admission (n=2). Moreover, twice as many of these infants (n=18) identified with a PPARC other than non-Hispanic white compared to those that identified as white (n=8).

We sought to assess the ability of neonatologists to predict which patients would receive a PrGD from the information available to them. We defined successful prediction of PrGD status as requesting a genetics consult in a patient who was later found to have a PrGD and not requesting a genetics consult in a patient who did not receive a PrGD. Overall, neonatologists requested a consult in 76% of patients with a PrGD (i.e. recall) and did not request a consult in 56% of patients without a PrGD (i.e. precision). However, we hypothesized that there were differences in their ability to predict a PrGD in non-white vs non-Hispanic white patients (Figure 5). We used a logistic model to assess the effects of PPARC, referral for a consult, and the interaction between these variables on prediction of a PrGD. Although the impact of neither PPARC nor whether a consult alone was significant, there was a significant interaction term (coef=1.8, p=0.034). This indicates non-Hispanic white infants referred for genetics consultation were 6.1 times more likely to receive a PrGD than non-white infants referred for a genetics consultation. Conversely, non-white infants who were not referred for a genetics consult were 2.4 times more likely to receive a PrGD than white infants who also were not referred. To confirm these findings, we used an alternative analysis approach assessing differences in precision (not referring patients who did not later receive a PrGD) and recall (referring patients who later did receive a PrGD). We observed relatively high precision (75%) and recall (86%) for non-Hispanic white patients while both precision and recall were markedly lower (55% and 68%, respectively) for non-white patients, yielding a composite F1 score of 0.61 for non-white patients vs. 0.8 for non-Hispanic white patients. We performed a bootstrapping analysis with 10,000 replicates to assess the significance of the differences in F1 score (p=0.013), precision (p=0.033), and recall (0.04) between non-white and non-Hispanic white PPARC groups.

**Figure 5.**
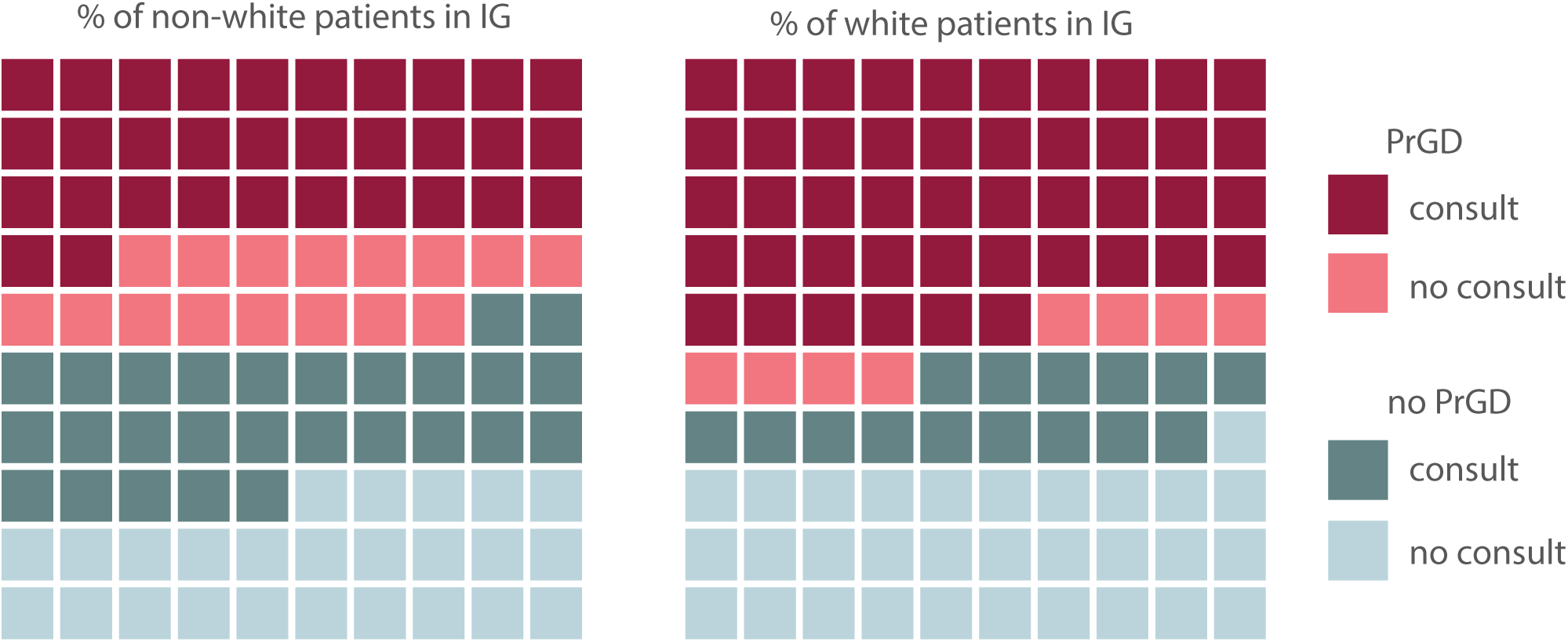
Impact of PPARC on prediction of precise genetic diagnosis in newborns. Prediction of precise genetic diagnosis (PrGD) status for non-white and non-Hispanic, white infants in the intervention group (IG) as assessed by patterns of referral for a genetics consult by their neonatologists. We defined successful prediction of PrGD status as requesting consultation on a patient who was later found to have a PrGD and not requesting consultation on a patient who was not found to have a PrGD. Neonatologists missed requesting a genetics consult significantly more often (2.4-fold) for non-white infants who would eventually receive a PrGD and also referred non-white infants for consults who were 0.17-fold less likely to go on to receive a PrGD. This translates to higher precision (75%) and recall (86%) for predicting PrGD status for non-Hispanic white infants than for non-white infants (55% vs. 68%, respectively).

Both analyses show that neonatologists were significantly less successful at predicting a potential PrGD in non-white patients compared to non-Hispanic white patients. That is they missed referring for genetics consult disproportionately more often in non-white patients who would eventually receive a PrGD and simultaneously referred more non-white patients for a consult who did not go on to receive a PrGD.

### Impact of a precise genetic diagnosis on changes in management

Using simple exclusion criteria to assess eligibility for testing and use of rWGS as a first-tier test revealed a high proportion of individuals across the IG with results that impacted their clinical care. These impacts fell in one or more of eight changes in management (COM) options (Figure 6). At 90 days after enrollment, access to a PrGD in the IG informed COM in 60 of 62 (97%) patients (Figure 5, Supplementary Table 9). Of the 60 families, anticipatory guidance was provided to 53 (88%) and 51 (85%) received recurrence risk counseling. Fifty-two families (87%) had at least one COM other than guidance or counseling. Of these families, the most common management change (38/52; 73%) was request for consultation from additional specialists for further evaluations, typically to facilitate medical decision-making, or consideration of diagnostic imaging procedures. In 16 (31%) of these patients, a PrGD resulted in request for additional laboratory testing, and a PrGD brought about a change in therapeutics in 10 patients (19%). In 15 of these 52 families (29%) with a change in management, a PrGD in their newborn had health-related implications for other family members leading to additional genetic testing and/or referrals for medical care (e.g. initiation of screening for hereditary hemorrhagic telangiectasia in a parent). Among 22 families of infants with a PrGD of a condition associated with early mortality, a PrGD contributed to a change in goals of care including a shift to palliative support in 5 (23%) infants.

**Figure 6.**
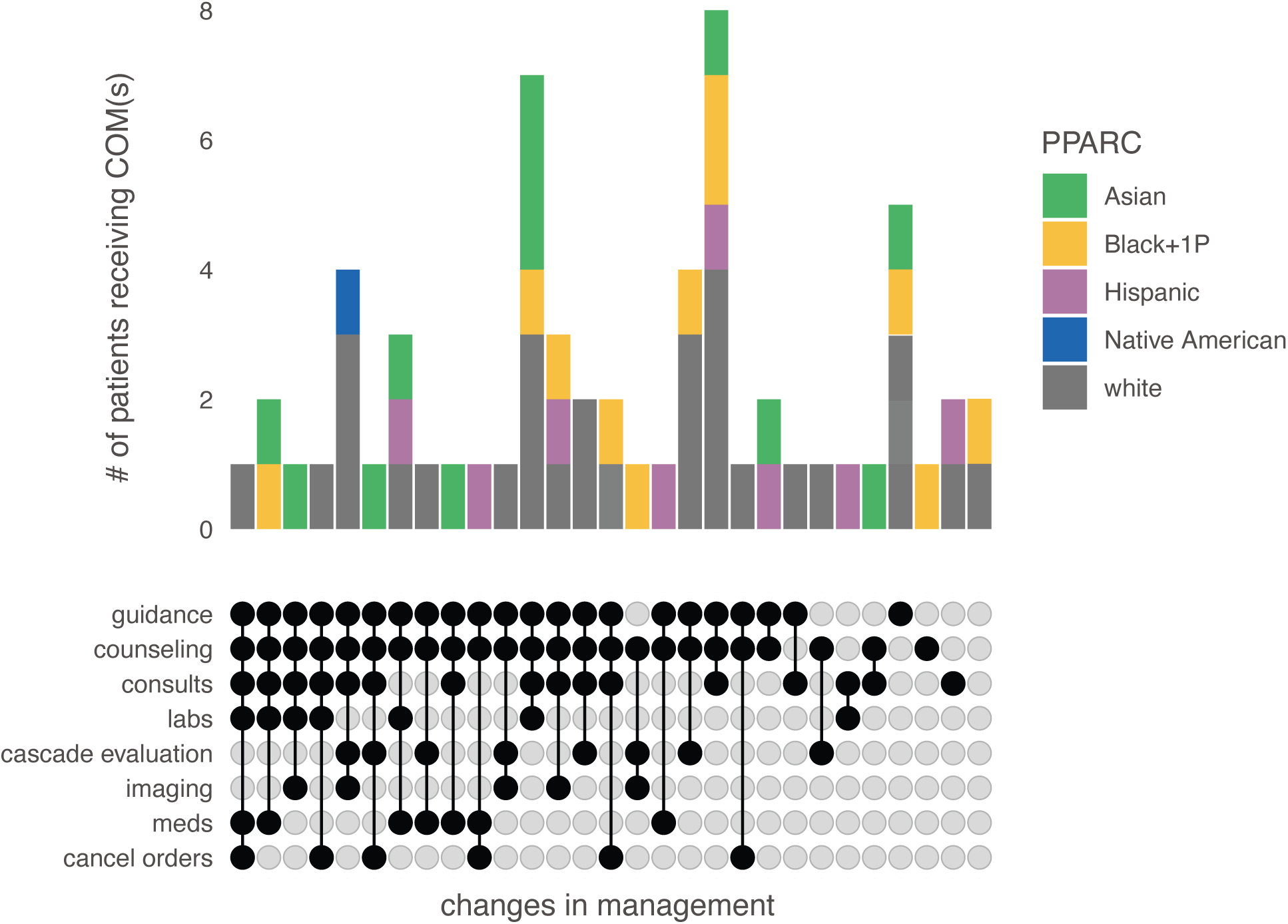
Impact of a precise genetic diagnosis on changes in management. The impact of a precise genetic diagnosis on changes in management (COM) stratified by parent or provider assigned racial construct (PPARC). Management options are reflected in the text at the lower left, with blackened circles representing assessment by clinical team. Multiple options could be selected. The vertical black lines connecting blackened circles represent a unique combination of COM options and all the unique COM combinations reported more than once are shown. Bar plots above each set of responses indicate the number of families with each COM combination stratified by PPARC. Abbreviations: anticipatory guidance (guidance), genetic counseling (counseling), request for additional consults (consults), ordering additional laboratory tests (labs), evaluation of other family members (cascade evaluation), ordering additional imaging studies (imaging), addition of medications (meds), canceling laboratory tests, imaging studies, or medications (cancel orders).

A total of 28 unique combinations of COM were observed, the most common of which included guidance, counseling, and request for consultation (Figure 5). We then tested whether, given a PrGD, a participant’s downstream care was impacted by comparing differences in the total number of COM and for any specific COM between non-Hispanic white and non-white PPARC groups and between non-Hispanic white versus Black+1P groups. None of the comparisons reached statistical significance (Wilcoxon rank sum test p > 0.2 for all comparisons). There was also no significant difference in total number of COM or any individual COM upon excluding anticipatory guidance and recurrence risk counseling.

## Discussion

We tested the impact of a genotype-driven workflow using simple, broad exclusion criteria to identify infants with critical illness who might benefit from rWGS as a first-line test, using access to a PrGD, diagnostic yield, access to a PrGD and diagnostic yield among PPARC groups, and missed PrGD in families not offered clinical genetic testing, as primary outcome measures. Use of this workflow significantly increased access to a PrGD and diagnostic yield in newborns admitted to the NICU and this increase was sustained over the ensuing year. Indeed, only 14% of infants who received conventional care were even offered ES by the age of fifteen months, suggesting that access to a PrGD further worsens upon discharge from the NICU, and only about half of these infants (6.1%) underwent ES because of declination or barriers to testing in the outpatient setting (e.g., insurance denials or requirement for additional clinical evaluations). In other words, not using exclusion criteria to determine eligibility for rWGS to infants in the NICU resulted in both an immediate and sustained loss of access to a PrGD and thereby multiple missed opportunities for intervention.

Significantly more infants identified as non-white or Black received a PrGD in the IG compared to the CCG, whereas access to a PrGD was comparable across all PPARC groups in the IG. In other words, access to a PrGD was equitable in the IG as a whole and across all PPARC groups compared to the CCG. This is due both to lack of access to genetic testing for infants in the CCG and that when testing was ordered for infants in the CCG, tests with lower diagnostic rates than rWGS were requested, particularly in non-white patients. Achieving equity in access to a PrGD will necessitate removing the many barriers that lead to disparities in underserved populations. Our results suggest that access to testing is a significant barrier in the NICU and that equitable access to a PrGD can be achieved by using simple exclusion criteria rather than conventional patient selection criteria as the basis for offering testing.

By comparing rWGS eligibility using simple, broad exclusion to typical inclusion criteria, we observed that the conventional workflow limits access to genetic testing and would have missed the opportunity to make a PrGD in 42% of infants with a PrGD in the IG, despite a substantial infrastructure for providing inpatient clinical genetics services. Most of these infants were non-dysmorphic with common clinical findings. Non-specific presentations of genetic diseases are common in the NICU, and some studies have suggested that requiring a suspicion of a genetic condition as a prerequisite to genetic testing will not only delay ordering testing but will miss up to half of all genetic conditions leading to NICU admission. With approximately 800 NICUs in the U.S. (most of which do not have access to clinical geneticists, genetic counselors or ES/WGS), and about 400,000 newborn admissions annually, tens of thousands of newborns admitted to NICUs likely have genetic conditions that are missed due to lack of access to genetic testing, much less access to a PrGD.^69^ Our approach mitigated disparities in access to genetic testing, particularly in non-whites.

A primary outcome metric used to assess the utility of ES / WGS, particularly in critical care settings, is diagnostic yield. We anticipated that by using a simplified clinical workflow that based eligibility on excluding infants whose clinical findings were fully explained by indications for NICU admission unlikely due to a genetic variant(s) with a large effect (i.e., prematurity, trauma, or infection), the diagnostic yield in the IG would be low in general and lower than that of standard care provided in parallel. Instead, the diagnostic yield in the IG (50%) was higher than in the conventional care group (26%). This is higher than the average diagnostic yield across more than thirty studies evaluating the use of rapid ES / WGS in pediatric critical care settings and accounted for largely by the PrGD made in infants who a neonatologist did not suspect to have a genetic condition based on clinical findings.

Use of diagnostic rate as a proxy for the value of genomic testing has, in our opinion, the potential to be misleading as a primary outcome measure. Diagnostic rates vary widely, often by several fold, across different cohorts, conditions, and clinical contexts, each of which can be defined in ways to maximize the pre-test probability of a PrGD and therefore diagnostic yield.^70^ But maximizing pre-test probability of a PrGD can adversely impact access to a PrGD. For example, the combination of subjectivity in case selection and the constraint of expert availability has motivated development and validation of objective criteria to stratify families for ES / WGS to maximize diagnostic rate. Such criteria are typically complex and challenging to navigate even for expert clinical geneticists. Moreover, use of such criteria can be labor-intensive, confusing, and intimidating for non-genetics providers (e.g., neonatologists). As a result of these complex clinical workflows and lack of support for non-genetics providers, critically ill infants who could benefit from genetic testing may often not be offered testing due to ineligibility against these criteria or never having been evaluated for eligibility in the first place. Simple exclusion criteria have the potential to improve access to a PrGD by offering a straightforward, objective workflow for clinicians.

Limitations of this study include that it was performed at a single site of care with potential differences in access to admission to the NICU and that its duration was limited to just a year. It is possible that differences in access to genetic testing and a PrGD between the IG and CCG are partly due to a selection bias resulting in more families in the CCG who were less interested in testing or a PrGD, and / or a higher number of deaths in the CCG such that testing could not be offered. However, of the 114 families enrolled in the CCG 90% (46/51) who were offered genetic testing consented to testing and while there were 10 deaths (9% of the cohort) in the CCG, 15 infants (12%) in the IG died in the first year of life. Differences in the presenting clinical findings between the IG and CCG might have also led to differences in access to testing and / or diagnostic yield. However, the percentage of patients with multiple congenital anomalies, isolated congenital anomalies, abnormal lab values, and seizure disorders was similar in each cohort. While survey and EMR data were available for the IG so that, for example, families could be directly questioned about the impact of a PrGD on clinical care, for the CCG, data were available only from the EMR. Lastly, the sample sizes of individual PPARC groups were small, necessitating labeling patients into broad categories such as non-Hispanic, white and non-white. This limits assessment of impact to a high, rather than granular, level thereby impeding identification of potential barriers to testing.

## Conclusions

Our results suggest that use of a simple workflow based on exclusion of only infants with clinical findings fully explained by prematurity, infection, or trauma to assess eligibility for rWGS coupled with support of provider readiness improves access to a PrGD and more equitable access to a PrGD. Major limitations of conventional workflows include dependence on use of complex stratification algorithms to maximize diagnostic rates and stepwise testing to minimize cost that alone, much less in combination, are challenging to operationalize and result in missed opportunities to make a PrGD, particularly in non-white infants. Collectively, our results demonstrate clear opportunities exist to improve equitable access to a PrGD in critically ill newborns and are a step toward establishing a practice to increase equity in the use of genomics in the critical care of infants and in Pediatrics in general.

## Supporting information

SeqFirst_Supplement

## Data Availability

All data produced in the present study are available upon reasonable request to the authors

## Acknowledgements

We thank the families for their participation and support. We thank the faculty and staff of the SCH NICU and the CICU. Financial support was provided by grants from GeneDx, the Brotman-Baty Institute, and University of Washington Center for Rare Disease Research (UW-CRDR) with additional contributions from NHGRI grants U01 HG011744, UM1 HG006493, and U24 HG011746. The content is solely the responsibility of the authors and does not necessarily represent the official views of the National Institutes of Health.

## Data and code availability

All data needed to evaluate the conclusions in the paper are present in the paper and/or the supplemental information. The rWGS data will be deposited in NHGRI’s Analysis Visualization and Informatics Lab-space (AnVIL) (accession number phs003047, “internal_project_id” contains seqfirst-neo). All other data are available upon request and if consistent with the informed consent of study participants.

