## Supplementary material for "SeqFirst: Building equity access to a precise genetic diagnosis in critically ill newborns": SeqFirst_Supplement

Order in Text:

Supplementary Table 1: Ineligible Participants (n=168)

Supplementary Table 2: Clinical Characteristics of Enrolled Infants

Supplementary Table 3: Declined Participation (n=83)

Supplementary Table 4: PrGD in CCG Group Offered Clinical Testing

Supplementary Table 5: PrGD Diagnostic Rate by Categorical Group

Supplementary Table 6: Conditions Diagnosed in the Intervention and CCG cohorts

Supplementary Table 7: PrGD Diagnostic Rate in Cardiac ICU (CICU)

Supplementary Table 8: Parallel Testing Offered to SeqFirst Group

Supplementary Table 9: Changes in Medical Management at 90 days based on PrGD

Supplementary Figure 1: Diagnoses Missed in Intervention Cohort by Conventional Workflow

| Supplementary Table 1: Ineligible Participants (n=168) | |
| --- | --- |
| **Exclusion criteria** | **Number excluded** |
| birth trauma | 64 (38%) |
| prematurity | 47 (28%) |
| infection | 36 (21%) |
| pre-existing PrGD | 15 (9%) |
| other | 6 (4%) |

| Supplementary Table 2: Clinical Characteristics of Enrolled Infants | | |
| --- | --- | --- |
| **Clinical features at presentation** | **Intervention (n=126)** | **CCG (n=114)** |
| multiple congenital anomalies (MCA) | 56 (44%) | 44 (39%) |
| MCA with CHD | 28 (22%) | 23 (20%) |
| MCA without CHD | 28 (22%) | 21 (18%) |
| isolated congenital anomaly | 33 (26%) | 33 (29%) |
| abnormal lab test | 10 (8%) | 11 (10%) |
| hematological abnormality | 2 (2%) | 1 (1%) |
| seizures | 9 (7%) | 5 (4%) |
| single or multi-organ failure | 6 (5%) | 12 (11%) |
| strokes | 3 (2%) | 1 (1%) |
| other | 7 (6%)* | 7 (6%)** |
| CHD=congenital heart disease *Thrombocytopenia, direct hyperbilirubinemia, anemia in setting of HLH, meconium ileus, complications of prematurity, chylothorax, possible lymphangiectasia, hydrops fetalis, thrombocytopenia, pelviectasis, right ventricular hypertrophy, encephalopathy, rhabdomyoma and Wolff-Parkinson-White syndrome, hypotonia and ectopic pituitary, congenital ichthyosis with collodion membrane **respiratory failure in setting of hydrops, persistent desaturations, renal artery thrombosis, stridor and poor feeding, recurrent SVT, SVT and apnea, hypotonia | | |

| Supplementary Table 3: Declined Participation (n=83) | |
| --- | --- |
| **Reason for Decline** | **Number Declined** |
| no response by deadline | 32 (39%) |
| feeling overwhelmed | 26 (31%) |
| belief that testing was unnecessary | 13 (16%) |
| uncomfortable with research | 7 (8%) |
| privacy concerns | 3 (4%) |
| concerns about blood draw | 2 (2%) |

| Supplementary Table 4: PrGD in CCG Offered Clinical Testing | | |
| --- | --- | --- |
| **Clinical genetic testing (by 15 months)** | **CCG (n=114)** | |
| exome | 7 (6%) | |
| any testing | 46 (40%) | |
| PrGD made from genetic testing | 12 (26%) | |
| **Parent or provider-assigned racial construct (PPARC)** | **CCG PrGD (n=12)** | **CCG no PrGD (n=102)** |
| Asian | 4 (33%) | 13 (13%) |
| Black | 0 (0%) | 12 (12%) |
| Hispanic | 2 (17%) | 18 (18%) |
| Native American | 0 (0%) | 3 (3%) |
| non-white, multi-racial | 0 (0%) | 7 (7%) |
| white, non-Hispanic | 6 (50%) | 42 (41%) |
| unknown | 0 (0%) | 7 (7%) |

| Supplementary Table 5: PrGD Diagnostic Rate by Categorical Group | | |
| --- | --- | --- |
| **Clinical features at presentation** | **Intervention (n=126)** | **CCG (n=114)** |
| multiple congenital anomalies (MCA) | 30/56 (54%) | 4/44 (9%) |
| MCA with CHD | 16/28 (57%) | 4/23 (17%) |
| MCA without CHD | 14/28 (50%) | 0/21 (0%) |
| isolated congenital anomaly | 11/33 (33%) | 2/33 (6%) |
| abnormal lab test | 7/10 (70%) | 1/11 (9%) |
| hematological abnormality | 2/2 (100%) | 0/1 (0%) |
| seizures | 5/9 (56%) | 2/5 (40%) |
| single or multi-organ failure | 3/6 (50%) | 1/12 (8%) |
| strokes | 0/3 (0%) | 0/1 (0%) |
| other | 5/7 (71%)* | 1/7 (14%)** |
| With CHDCHD=congenital heart disease *Thrombocytopenia, direct hyperbilirubinemia, anemia in setting of HLH, meconium ileus, complications of prematurity, chylothorax, possible lymphangiectasia, hydrops fetalis, thrombocytopenia, pelviectasis, right ventricular hypertrophy, encephalopathy, rhabdomyoma and Wolff-Parkinson-White syndrome, hypotonia and ectopic pituitary, congenital ichthyosis with collodion membrane **respiratory failure in setting of hydrops, persistent desaturations, renal artery thrombosis, stridor and poor feeding, recurrent SVT, SVT and apnea, hypotonia | | |

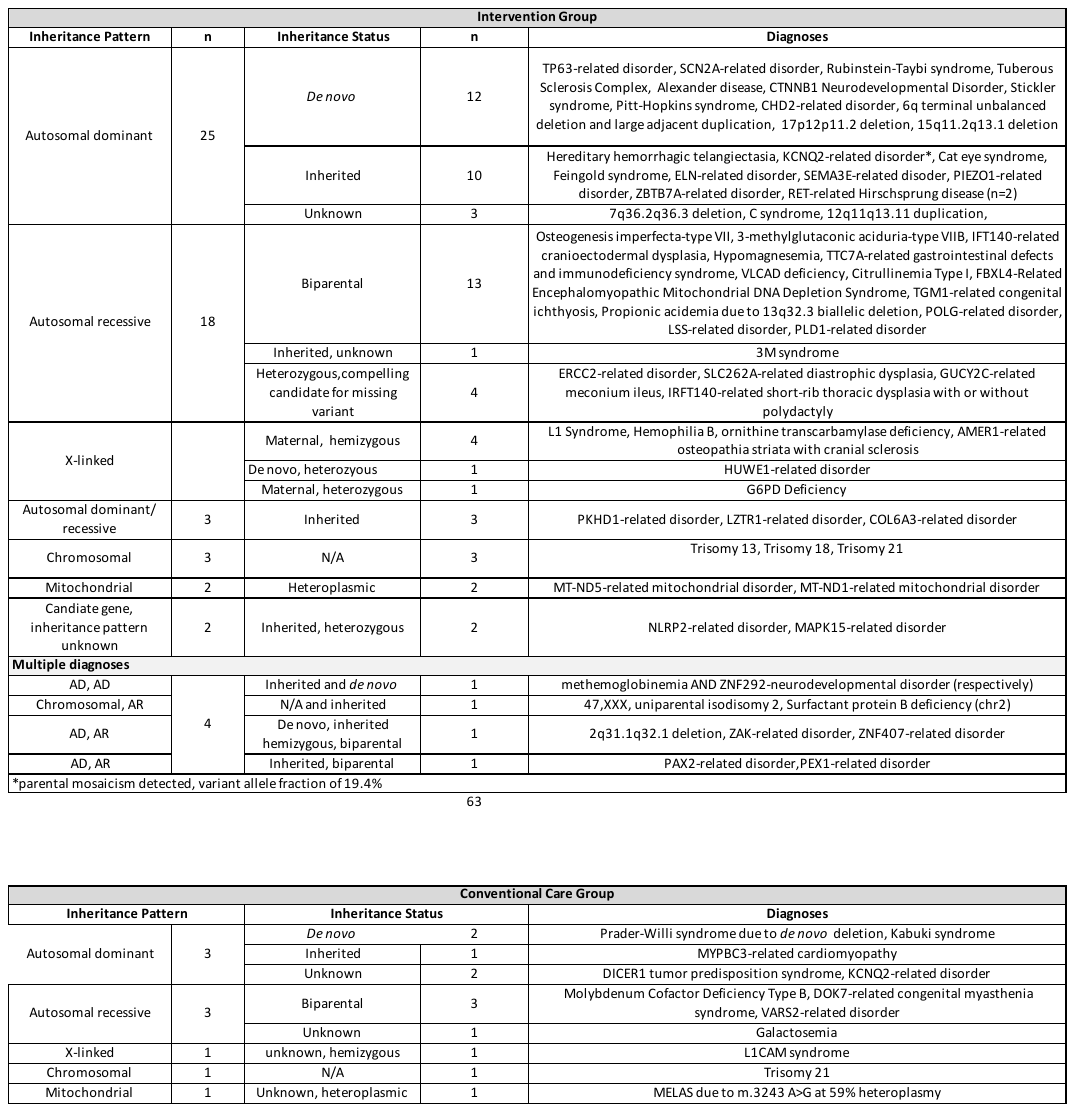

Supplementary Table 6: Conditions Diagnosed in the Intervention and CCG cohorts

| Supplementary Table 7: PrGD Diagnostic Rate in Cardiac ICU (CICU) | |
| --- | --- |
| **Clinical features at presentation** | **Intervention Group CICU (n=25)** |
| isolated congenital heart disease (CHD) | 3/15 (20%) |
| CHD + other congenital anomalies | 5/9 (56%) |
| arrythmia | 0/1 (0%) |
| **Total** | **8/25 (32%)** |

| Supplementary Table 8: Parallel Testing Offered to Intervention Group | |
| --- | --- |
|  | **Intervention PrGD (n=62)** |
| **No genetics consult*** | 15/62 (24%) |
| "non-dysmorphic" features | 14/15 (93%) |
| non-white PPARC group | 10/15 (67%) |
| **Suspicion for genetic condition (i.e. consult ordered)** | 11/62 (18%) |
| genetic testing completed, but non-diagnostic | 9/11 (82%) |
| infant demise prior to clinical genetic testing | 2/11 (18%) |
| **Missed by conventional workflow** | 26/62 (42%) |
| ***Diagnoses made with no suspicion of genetic condition** |  |
| Mitochondrial disease (n=2) |  |
| *RET*-related Hirschsprung Disease (n=2) |  |
| *ACVRL1*-related Hereditary hemorrhagic telangiectasia |  |
| *CHD2*-related disorder |  |
| *COL6A3*-related myopathy |  |
| *ELN*-related pulmonary stenosis |  |
| *F9*-related hemophilia |  |
| *GUCY2C*-related meconium ileus |  |
| *KCNQ2-*related epilepsy |  |
| *MAPK15*-related disorder |  |
| *MYCN*-related Feingold syndrome |  |
| *CREBBP*-related Rubenstein-Taybi syndrome |  |
| *TTC7A*-related disorder |  |

Supplementary Table 9: Changes in Medical Management at 90 days based on PrGD

| **Medical Management Changes (COM)** | **Intervention group all (n=62)** | **Intervention group with COM (n=60)** |
| --- | --- | --- |
| anticipatory guidance | 53 (86%) | 53 (88%) |
| risk counseling | 51 (82%) | 51 (85%) |
| new consult(s) | 38 (61%) | 38 (63%) |
| new lab tests | 16 (26%) | 16 (27%) |
| cascade evaluation of family members | 15 (24%) | 15 (25%) |
| new imaging | 10 (16%) | 10 (17%) |
| changes in medication | 10 (16%) | 10 (17%) |
| cancel orders | 7 (11%) | 7 (12%) |

| **Any medical management changes** | **60/62 (97%)** |
| --- | --- |

Supplementary Figure 1: Diagnoses Missed in Intervention Cohort by Conventional Workflow

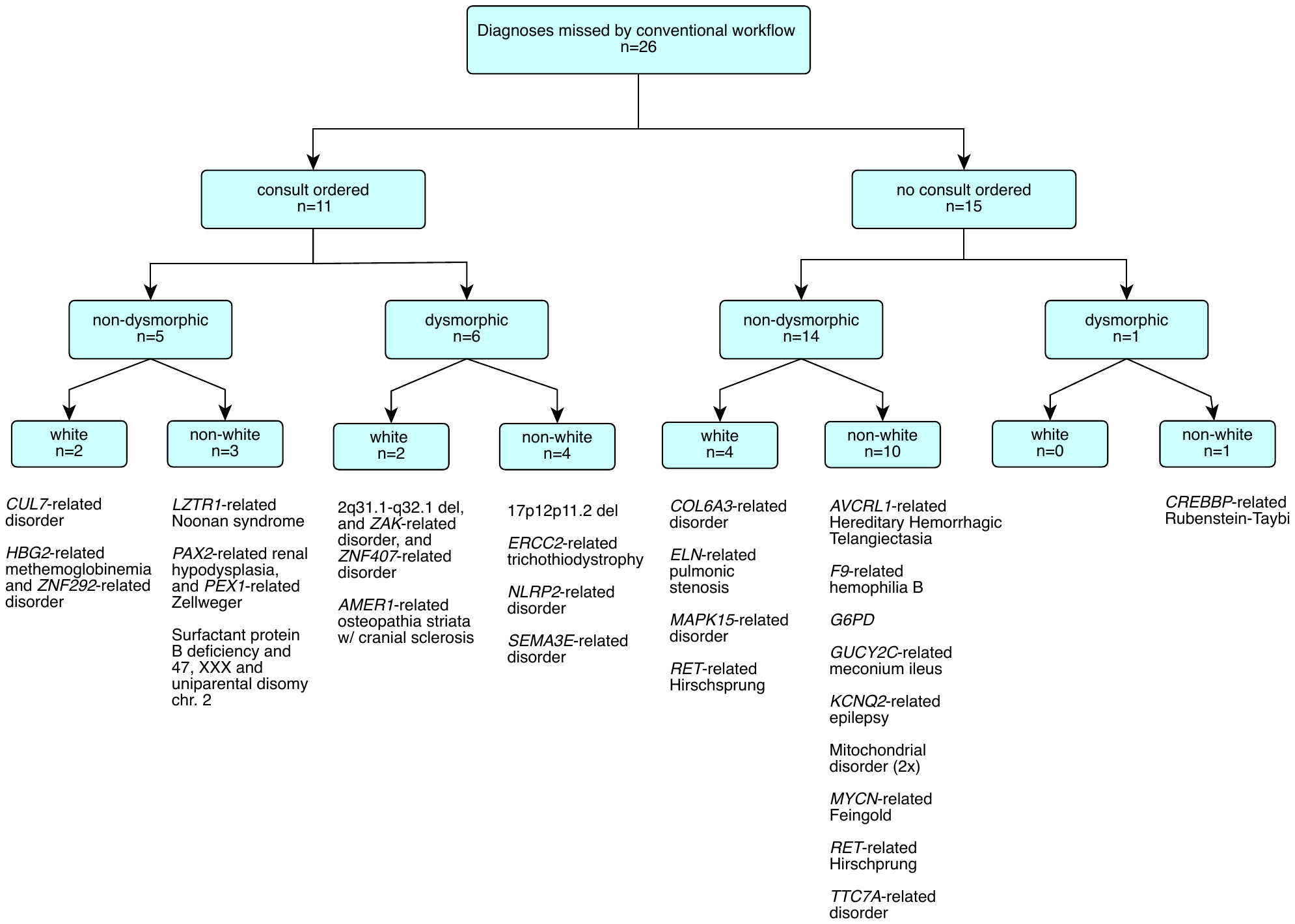
